# Detection of lung hypoperfusion in Covid-19 patients during recovery by digital imaging quantification

**DOI:** 10.1101/2020.05.29.20117143

**Authors:** Gianluigi Patelli, Silvia Paganoni, Francesca Besana, Fabiana Codazzi, Mattia Ronzoni, Simone Manini, Andrea Remuzzi

## Abstract

**Purpose:** A large number of patients affected by the SARS-Cov-2 virus worldwide undergo recovery of symptoms in about one month. Among these patients, the healing process is still under observation, with some patients in need of careful clinical monitoring. While the radiological findings have been shown to regress almost completely, little knowledge is available at the moment about other complications in the lung and in other organs. We then investigated the lung perfusion conditions in patients affected by COVID-19 during recovery.

**Method:** We retrospectively studied 20 patients, from 14 to 60 days after resolution of the COVID-19 symptoms, using chest CT. In a subgroup of 5 patients contrasted CT was used. Beside normal radiological evaluation of lung tissue, perfusion conditions were evaluated by digital image processing in the lung volume automatically segmented.

**Results:** Pulmonary lung evaluation showed that COVID-19 pneumonia almost completely regressed, with mild focal areas affected by fibrous stripes. In patients that reported dyspnea, lung CT showed complete resolution of interstitial changes. Quantification of lung perfusion condition by contrasted CT, showed that dyspnea in 3 patients was associated with areas of hypoperfusion, while in 2 patients not reporting dyspnea perfusion conditions were comparable to normal controls.

**Conclusions:** Although we obtained preliminary data, this is the first report on quantitative evaluation of hypoperfused lung tissue detected in recovering COVID-19 patients. These results suggest the need to further investigate these patients and to redefine the role of CT evaluation for diagnostic purposes as well as for evaluation of potential treatments.

**Funding:** This was an academic study that received no direct funding.

## Introduction

The pathophysiology of SARS-CoV2 is becoming clearer with a number of recent reports in the literature after the fast viral outbreak in the city of Wuhan in China and more recently in other countries worldwide[1]. The major symptom of SARS-CoV2 is severe acute respiratory syndrome due to virus interaction with cells expressing angiotensin-converting enzyme 2 (ACE2) and TMPRSS2, such as epithelial cells, endothelial cells, alveolar macrophages, triggering important immune response with the generation of cytokines and chemokines, which attract monocytes, macrophages and T cells promoting further inflammation[2]. This may lead to further accumulation of immune cells in the lungs, with overproduction of pro-inflammatory cytokines and activation of complement and coagulation systems in lung microcirculation[3,4]. During the course of the disease, this important immune response attracts virus-specific T cells to the site of infection, where they can eliminate the infected cells. Alveolar macrophages can then recognize neutralized viruses and apoptotic cells and clear them by phagocytosis. Altogether, these processes lead to clearance of the virus and minimal lung and other organ damage, resulting in recovery[5].

Despite these pathophysiological mechanisms are under continuous investigation, and the results of clinical studies presented in the literature on the effects of different therapeutical approaches on the SARS-CoV2 infection, there are only a few reports on the clinical course during the recovery phase of the disease. Recent investigations[6,7] show that lung abnormalities on chest computerized tomography (CT) show the greatest severity approximately 6–11 days after the initial onset of symptoms, while in the following weeks’ recovery occurs consistently and in around 30 days lung tissue lesions are almost absorbed[8]. However, extended observation on COVID-19 patient during recovery are still object of investigation, with and without positive result for RT-PCR test.

Due to the large number of patients admitted to the Bolognini Hospital (ASST Bergamo Est company) in the Bergamo area, during the COVID-19 outbreak, we had the chance to review CT findings in a number of patients during the recovery from the disease. In particular in some of these patients, CT scans were used to investigate potential thrombotic events with and without the use of contrast media. We report here the results of these investigations, since the use of the contrast media allowed us to investigate the perfusion conditions of these organs using digital image processing. We did identify perfusion defects in lungs of these patients, more than one month after remission of the symptoms, that deserve attention.

## Material and Methods

### Patient population

During the period from 15 April and 30 April, 20 patients (12 male and 8 female; age range 35–86, mean age 58 ±10 years) previously treated for pneumonia SARS-CoV-2, with negative swab, have been admitted to our department to assess the outcome of SARS-CoV-2 pneumonia. The follow up was performed by chest CT at an interval between 14 days and 60 days after remission of the fever (average 40 ±13 days). Twelve patients reported an almost complete resolution of the symptoms, while 8 patients reported residual dyspnea. Of these 8 patients, 3 reported dyspnea with minimal motor activity, while the others after prolonged effort. Control CT was performed, without contrast media in 15 patients and with contrast media for suspected thromboembolism in 5 patients (including those with dyspnea with minimal motor activity, or patients with D-Dimer value over normal range). Four patients not affected by COVID-19, studied by angioCT scan for other clinical indications and with normal lung tissue, were used as a control group. Ethical approval of this retrospective evaluation was obtained by the Bergamo province Ethics Committee (Reg. Sperim. N. 80/20 on 22/04/2020).

### Chest CT acquisitions

Conventional chest CT (CT 128 slice Ingenuity, Philips, Amsterdam, The Netherlands) was performed with the patient in the supine position during end-inspiration. The chest CT protocol used is as follows: from apices to mid-renal; slice thickness 1mm; slice increment 1mm; pitch 0.94; Rotation time 0.5 sec; Field of view 411 mm; Voltage 120 kV; mAs modulation 100–200 mA. The acquisition protocol consisted of not contrasted acquisition, arterial phase acquisition (80 mL/Iomeron 400, injection 4 ml/sec; Threshold 90 HU), venous phase acquisition (60 sec from the threshold). Contrast media was infused through the antecubital vein with a 18- or 20-gauge catheter. Images were viewed on a PACS monitor using IMPAX version 6.6.1 (Agfa-Gevaert NV Septestraat 27B-2640 Mortsel – Belgium). Every chest CT examination was evaluated by two double-blind radiologists with many years of experience in interpreting chest CT.

The lung perfusion evaluation was performed by using PAA (Pulmonary Artery Analysis) Software installed on IntelliSpace Portal release 11 (Philips Medical Systems, Best, the Netherlands)[9]. PAA provides semi-automatic and manual tools to visualize and measure pulmonary embolism and provides a Hounsfield Units (HU)-based colormap visualization tool. In particular the default setting allows imaging tissue lung perfusion using color palette for HU ranging from −749 to −983. The threshold level identified for normal lung tissue perfusion in control subjects was in average equal to −890 HU. Lower attenuation values can be considered areas of hypoperfusion. In order to precisely quantify the volume of lung tissue identified as low perfusion, independently by the amount of contrast media, we used the following method to assume the HU threshold value discriminating between normal and hypoperfused tissue. We calculated, that the previously mentioned range of HU for hypoperfused tissue (HU lower than –890) in the control group was on average 13.3% of the total HU range, estimated from air to pulmonary artery. Assuming this percentage, we calculated the new threshold level for hypoperfused tissue at the single patient level, using estimation of the HU range from air to pulmonary artery, and considering 13.3% of this interval, starting from HU of the air (−749). We labeled each pixel inside the lung mask as low perfusion or normal perfusion based on these patient-specific thresholds using Python implementation of SimpleITK library(*https://simpleitk.org/about.html*) to operate on DICOM images. To estimate the percentage of lung volume characterized by hypoperfusion, we initially segmented the lung volume of the CT scans by automatic lung segmentation using the u-net (R231) convolutional network[10]. This model was trained on a large dataset, including COVID-19 CT slices, that covers a wide range of visual variability. Segmentation on individual slices allowed to extract the right and left lung mask separately. The trachea was not included in the lung segmentation. The number of voxels with HU value in the range of hypoperfused tissue was visualized using DICOM Vision (*www.dicom.vision*) and the ratio of this voxel count over that of the total lung volume was calculated.

## Results

Out of the 20 patients who underwent follow-up, 12 patients had almost complete resolution of symptoms, with complete regression of the thickening areas detected with chest CT scan at diagnosis of SARS-CoV-2 pneumonia in 4 patients, and persistence of fibrous stripes areas in the other 8 patients. In the remaining 8 patients, 5 reported dyspnea after prolonged effort, with 3 of them with complete remission of signs of interstitial pneumonia and 2 with only partial remission. While the 3 other patients reported dyspnea with minimal effort and showed a completely normal CT scan and remission of parenchymal opacity. Out of the total number of 20 patients, 5 patients have been studied with contrast media. Three of them were those that reported dyspnea with minimal effort, while 2 had no such symptom. At CT evaluation, in all these patients no signs of thromboembolism were present. At variance, in the three patients affected by dyspnea color map representation, as shown in Figure 1, showed diffuse signs of hypoperfused areas (dark red/violet color) and the distribution of these areas was not uniform between the left and right lung. Quantification of hypoperfused volumes are reported in Table 1. Lungs from 3 out of 5 COVID-19 patients exceeded this threshold, reaching in one patient about 20% and 30% in the left and right lung. Of interest, tissue lung perfusion was normal in the two patients without dyspnea (see Table 1 and Figure 1). No evident sign of hypoperfused regions were present in CT of normal controls, and the hypoperfused tissue was present in less than 7% of lung volume.

**Table 1.** Percentage of lung volume occupied by hypoperfused tissue.

| Subject | Left Lung | Right Lung | Time after symptoms remission (days) | Patient symptoms |
| --- | --- | --- | --- | --- |
| Ctrl 1 | 4.39 % | 3.84 % | - |  |
| Ctrl 2 | 0.09 % | 0.35 % | - |  |
| Ctrl 3 | 6.20 % | 5.12 % | - |  |
| Ctrl 4 | 7.26 % | 6.58 % | - |  |
| Pt 1 | 19.05 % | 29.34 % | 54 | Dyspnea ++ |
| Pt 2 | 15.40 % | 11.01 % | 30 | Dyspnea ++ |
| Pt 3 | 13.48 % | 7.51 % | 30 | Dyspnea ++ |
| Pt 4 | 4.94% | 2.43% | 25 | - |
| Pt 5 | 3.73% | 4.75% | 36 | - |

**Figure 1:**
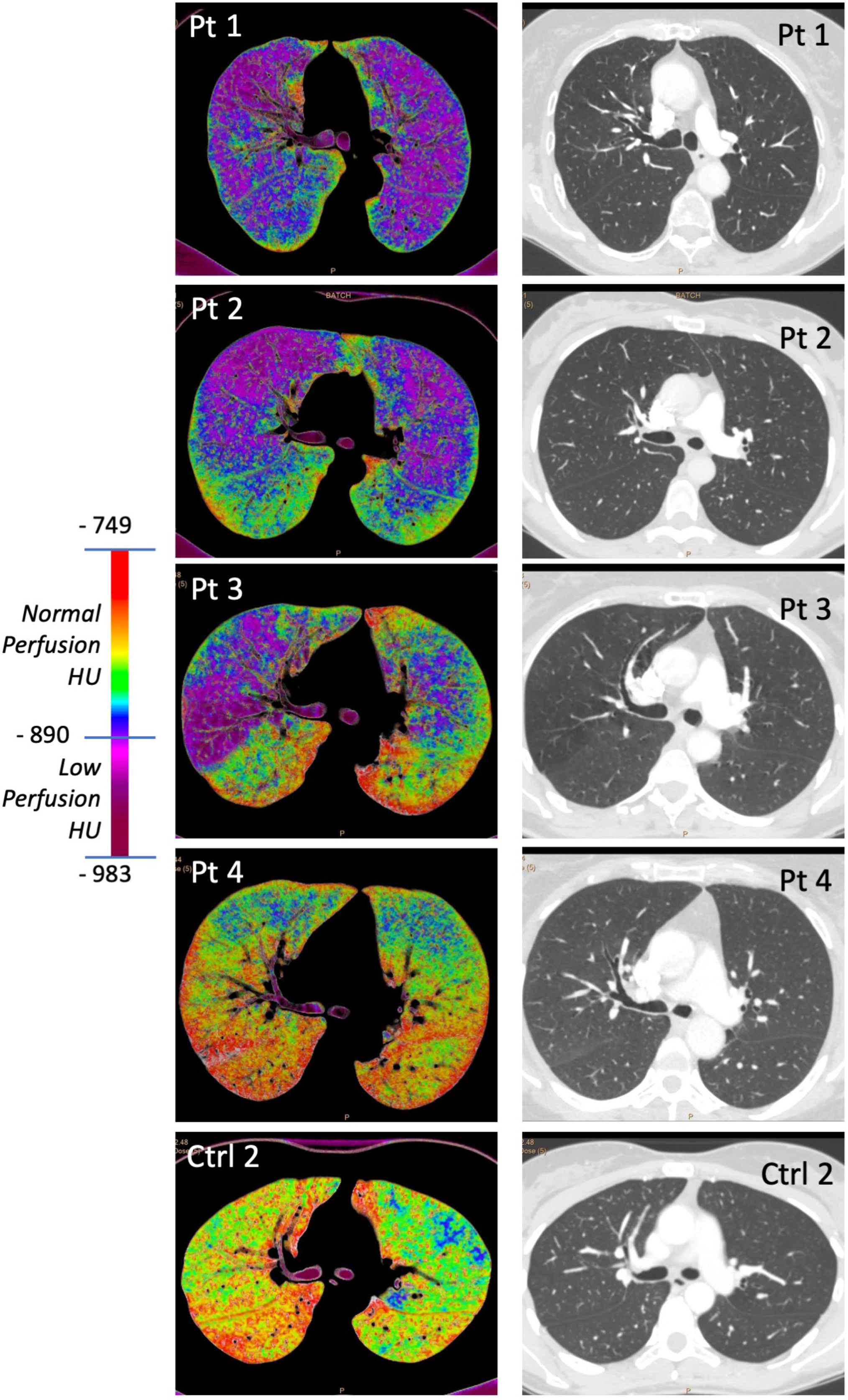
Contrast and non-contrast enhanced CT in COVID-19 patients and in normal control. Representative images of lung perfusion map and CT in COVID-19 patients (Pt1–4) and in normal control (Ctrl2). Perfusion maps of Pt1, 2 and 3 that were not affected by dyspnea show areas of low perfusion, while Pt4 that was not affected by dyspnea shows normal perfusion map, comparable to normal control (Ctrl2). Pulmonary CT was normal in all CT scans.

## Discussion

As mentioned previously, the lung tissue inflammation induced in COVID-19 patients by accumulation of immune cells, overproduction of pro-inflammatory cytokines and activation of complement and coagulation system in the microcirculation is the reason for the observed focal or diffuse damage to the lungs, as observed during the course of the disease. This acute lung damage correlates with the initial appearance of areas of increased density with the “ground glass” appearance, which gradually tends to change into areas of consolidation, preferentially located in the peripheral subpleural area. Four stages of infection were recently proposed: early, progression, peak, and resolution[11]. In the later stages of the infection, the pattern referred to as “crazy paving” and “reversed halo sign” have been found more frequently[11].

Our retrospective analysis, in negative RT-PCR test patients recovering from COVID-19, more than 60% of patients without symptoms chest CT, one month after the resolution of the disease, displayed an incomplete regression of the lung opacities, with the persistence of mild fibrous stripes in interstitial areas, while in patients who reported residual dyspnea, an almost complete regression of the parenchymal thickening was observed. At variance, our quantitative evaluations by volumetric image processing, show that in symptomatic patients (dyspnea), despite the absence of pulmonary fibrous stripes residues, there were hypoperfused areas of lung parenchyma, not symmetrical in the two lungs. These findings suggest that during the recovery of COVID-19 rather than ventilatory dysfunction, defect in lung blood perfusion of the microcirculation may persist.

To our knowledge, this is the first report on quantitative estimation of lung perfusion indicating the presence of microvasculature defect in recovering COVID-19 patients, without thromboembolism, and with normal pulmonary ventilation. Our results suggest that in COVID-19 patients incomplete healing of alveolar parenchyma microcirculation may occur or persist during recovery. Thus, while the lung tissue damage related to interstitial fluid accumulation and impairment of ventilation is almost completely recovered within one month, a persistent defect of the microcirculation may remain, likely due to residual of viral-induced inflammation, with immune cell accumulation, platelet adhesion and micro-disseminated thrombi. Whether also fibrosis may be responsible for these microcirculation perfusion defects is worth to investigate. While these complications of the disease are now well recognized during the acute phase of the infection[12,13,14], it is important to notice that this impairment of the microcirculation, we have detected, may affect lung function even in the recovery phase and may last for a long time or even not be healed with time. This evidence should be carefully considered, due to the potential clinical relevance of the problem in the large number of patients affected by the SARS-CoV-2 infection during the recovery phase worldwide in this period.

## Conclusions

In summary, the first message from our observation is that these patients, if still reporting dyspnea during recovery, should be followed using chest CT, to identify and quantify the presence of lung perfusion dysfunction. This will be important not only for diagnosis, but also to determine whether this damage to the lung microcirculation resolves with time or if they will be chronically affected by these changes. The second message of our observation is that the study of these patients may allow identification of the potential need for pharmacological interventions, to choose the correct drug treatment to adopt, and the duration for these treatments. Deeper knowledge of these pathological processes will also improve the clinical outcome of this large patient population that is still increasing in size due to the ongoing SARS-CoV-2 outbreak. While more extensive investigation with CT in discharged COVID19 patients is progress by our center and by others, we believe it is urgent to draw attention to these lung complication.

## Data Availability

Data are available upon request to the corresponding Author.

## Contributors

GP, SP, FB and FC performed data collection, analysis and clinical observations, MR and SM implemented computational codes and performed image processing, AR analyzed the data, AR and GP interpreted the data and wrote the manuscript. All authors interpreted the findings, contributed to manuscript revision, and approved the final version for publication.

